# Feasibility, acceptability, and appropriateness of the *Familia Bora* parenting program for couples with young children in Mwanza, Tanzania: a pilot mixed-methods process evaluation

**DOI:** 10.1101/2025.09.16.25334267

**Authors:** Joshua Jeong, Valencia J. Lambert, Carina Wolfram, Malia Uyehara, Gabriel Sangarara, Julieth Joseph, Megan Louttit, Juliet McCann, Damas Joachim

## Abstract

Through a research-practice partnership with collaborators and community members in Tanzania, we co-designed *Familia Bora*, a 17-session father-inclusive parenting intervention delivered weekly to couples in mixed-gender groups to promote responsive caregiving, couples’ relationships, caregiver mental health, and gender equity and holistically improve early child development. From July to November 2024, we piloted *Familia Bora* and conducted a mixed-methods process evaluation to assess its feasibility, acceptability, and appropriateness, and to identify opportunities for strengthening program delivery and implementation. Sixty-two couples with a child under 2 years were enrolled into the program across six communities in Ilemela District, Mwanza, Tanzania, from July to November 2024. Feasibility, acceptability, and appropriateness were assessed using multiple data sources: 90 session attendance records, 75 structured session observations, 79 facilitator feedback forms, 167 participant exit interviews (68 mothers and 99 fathers), endline surveys with 58 fathers, and 37 qualitative interviews at midline and 47 at endline with a purposively selected subsample of mothers, fathers, and group leaders. Quantitative data were analyzed descriptively, and qualitative data were analyzed using thematic content analysis. Findings were triangulated across data sources to provide a comprehensive assessment of program implementation across multiple dimensions. We found that the program was feasible to deliver and highly acceptable to both fathers and mothers. Attendance rates were satisfactory among both mothers and fathers. The majority of participants rated the program positively, highlighting its content relevance, cultural appropriateness, and perceived benefits. Sessions on responsive caregiving, economic strengthening, and mental health were rated most favorably, whereas topics like non-violent discipline and fathers’ engagement in household chores met some initial resistance due to gender and cultural norms. Parents valued the mixed-gender group format, interactive materials, and participatory delivery, noting that these aspects fostered learning, reflection, and message sharing within their communities. Transport reimbursements were also valued by participants and identified as a key facilitator of attendance. Overall, this pilot evaluation demonstrates that *Familia Bora* was feasible, highly acceptable, and well received by both mothers and fathers. Findings highlight the promise of co-designed, gender-transformative, couples-based parenting programs for reducing barriers, promoting equitable caregiving, and fostering supportive caregiving environments for early child development. These findings support further testing of this program through a larger implementation study in Tanzania and potentially other similar contexts.

## INTRODUCTION

Parenting interventions can effectively improve early child development (ECD) and strengthen parenting outcomes across diverse global contexts [1]. In low-resource settings in particular, a range of program models and delivery strategies have been developed, tested, and successfully implemented to expand parenting support and promote ECD [2]. Although most programs have primarily focused on mothers, there is growing recognition of the importance of actively engaging fathers in parenting interventions [3, 4]. Systematic reviews indicate that father-inclusive parenting programs can achieve wide-ranging benefits across the family, including on paternal and maternal parenting, couples’ relationships, gender dynamics, and young children’s health, development, and wellbeing [5].

As programmatic efforts to engage fathers have increased, various delivery models and implementation strategies have been explored [6]. In many cases, fathers are invited to join interventions originally designed for mothers and often at only a few selected sessions [7], while fewer programs engage men consistently as direct beneficiaries throughout [8]. Program models also vary in group composition, with some having mixed-gender sessions where mothers and fathers participate together throughout [9, 10], whereas others engage fathers mostly in gender-specific groups separate from mothers [11, 12]. These variations reflect differences in contextual conditions, programmatic goals, and even underlying assumptions. However, the rationale for program design decisions regarding father engagement is rarely articulated, with limited monitoring and evaluation (M&E) evidence to inform the most effective strategies in a given context.

Emerging evidence suggests that programs intentionally designed for fathers from the outset and through applying implementation science methods such as pretesting, refining, and piloting strategies are more likely to be effective and to sustain men’s interest and engagement [13]. Factors such as program content, how the intervention is framed, who delivers it, and whether men see their roles and experiences reflected in the program approach all influence fathers’ participation and engagement [6, 14]. At the same time, contextual barriers such as restrictive gender norms and the opportunity costs men face as financial providers of the family consistently undermine fathers’ engagement and the success of these programs across settings [15, 16]. Thus, programs that are responsive to men’s lived realities and cultural expectations are more likely to be seen as relevant, acceptable, and engaging. Yet few parenting interventions have been systematically designed and implemented to jointly address the needs and preferences of both mothers and fathers, especially in low-resource settings.

To address this gap, we co-designed *Familia Bora*, a multicomponent parenting intervention for couples with young children in Mwanza, Tanzania [17]. From 2022 to 2024, we developed this program through a rigorous six-step process that combined formative research, iterative co-design with fathers and mothers, and rapid pretesting cycles of each individual session across 13 communities. Ultimately, *Familia Bora* embraces a holistic, gender-transformative, and developmental approach and aims to strengthen parenting practices of both mothers and fathers, enhance couples’ relationships, address gender norms, and support caregiver mental health, to eventually in turn improve ECD.

Following this development phase, we conducted a pilot study in 2024 to evaluate the implementation of *Familia Bora* when delivered in its entirety to mothers and fathers. This process evaluation aimed to assess whether the program’s father-inclusive design achieved its goals and effectively addressed the needs and preferences of both parents. The specific objectives of this current study were to examine the feasibility, acceptability, and appropriateness of *Familia Bora* among mothers and fathers. By combining qualitative and quantitative methods and data sources, this mixed-methods evaluation sought to identify core components that worked well and areas for refinement to strengthen delivery and support larger-scale implementation and evaluation in Tanzania and similar settings.

## METHODS

### Setting and Context

This study was implemented in Ilemela District, a peri-urban area of Mwanza region, Tanzania. As of 2022, the region had a total population of 3,699,872, of which 509,687 resided in Ilemela District [18]. The primary economic activity in the region is agriculture followed by fishing, industrial processing of fish for export markets, and large-scale gold and diamond mining activities in neighboring regions [19].

### Study design

The overall goal of this pilot study was to evaluate Familia Bora, a newly developed parenting intervention designed to improve parenting practices, couples’ relationship dynamics, caregiver mental health, gender equity, and ECD outcomes. We used a pre-experimental evaluation design to explore preliminary pre-post changes on targeted outcomes and embedded a mixed-methods process evaluation to assess intervention feasibility, acceptability, and appropriateness. Findings were intended to assess proof of concept and inform curriculum revisions and key decisions to the program model in preparation for a larger cluster-randomized controlled trial. This paper specifically focuses on the process evaluation component, drawing on qualitative and quantitative data to examine how the intervention was delivered, how it was experienced by participants, and its perceived relevance and value from the perspective of both fathers and mothers. A separate manuscript is in progress that will report changes in father-, mother-, and child-level outcomes.

This study was conducted in collaboration with the Tanzania Home Economics Organization (TAHEA-Mwanza), a local non-governmental organization focused on parenting and ECD. In consultation with village chairpersons, six communities within Ilemela Municipal were selected for the study. In each community, the chairperson compiled a list of eligible households, from which TAHEA-Mwanza randomly selected 10 households in each village using simple random sampling. Caregivers were invited to participate in the parenting group sessions based on whether they met the following eligibility criteria: (1) primary male/female caregiver aged 18-65 years, (2) has a child younger than 2 years of age, (3) is in a current relationship with the child’s female/male caregiver, (4) has lived in the same household as their partner and index child during the past month, and (5) provided written informed consent to participate in the study.

### Intervention Description

Familia Bora is a group-based parenting intervention designed to improve parenting practices, couples’ relationship dynamics, parental mental health, and gender norms to ultimately improve ECD outcomes. The pilot intervention consisted of 15 sessions that were held on a weekly basis (approximately 4 months total from July to November 2024) at a central community location and at a time that was preferred by the majority of participants in each community. Overall, the full curriculum addressed a wide range of topics, including early childhood development, responsive parenting, positive discipline, couples’ communication, equitable decision-making, conflict resolution, intimate partner violence (IPV), caregiver mental health, economic strengthening, and nutrition. Thirteen of the 15 sessions were attended by couples with their children, while two sessions were designed specifically for fathers. The father-only sessions focused on the meaning of fatherhood and male caregivers’ mental health. Further details on the iterative and community-engaged process used to co-develop Familia Bora are available elsewhere [17].

Sessions were delivered in Kiswahili by a trained staff member from TAHEA-Mwanza, who had expertise in parenting, ECD, nutrition, and facilitating community-based programs. Each group also elected a male group leader to help mobilize participants and serve as a liaison between caregivers and the session facilitator from TAHEA-Mwanza. Facilitators followed a manualized curriculum using structured session guides that were designed with a consistent format that included a recap of the previous session, key lessons, interactive activities, and concluded with a short homework assignment. Sessions incorporated a range of behavior change techniques (BCTs) to support caregiver learning and engagement, including group discussions, problem-solving, demonstrations, guided practice, and goal setting. In addition to the facilitator’s guide, various other materials were incorporated throughout the sessions, including flipcharts with illustrative images, a tablet used for showing videos of early learning and responsive caregiving, toys and play materials for children to use during sessions, worksheets for small group activities, a play and communication guide with suggested activities for caregivers and a printed one-page handout with key illustrations and the homework assignment that was distributed after each session.

### Data collection

Data were collected from multiple sources to capture diverse perspectives on the process and quality of program implementation, focusing on the broad dimensions of feasibility, acceptability, and appropriateness. Drawing on established definitions from implementation science [20, 21], we defined *feasibility* as the extent to which the program was delivered as intended; *acceptability* as participants’ satisfaction with the content and delivery; and *appropriateness* as the perceived relevance and cultural fit in the given practice setting. These dimensions of process and implementation were assessed across various forms, respondents, timeframes, and data sources.

**Table 1** summarizes the seven sources of data collected in our process evaluation: attendance forms, participant dropout forms, M&E observation forms, facilitator feedback forms, participant feedback forms, endline quantitative surveys, and the midline and endline qualitative in-depth interviews. The table also indicates which process evaluation dimensions – feasibility, acceptability, and/or appropriateness – were assessed by each data source based on the types of questions or observations included in each form. Altogether, the process evaluation drew on 90 session attendance records, 2 dropout forms, 75 structured observations, 79 facilitator feedback forms, 167 participant exit interviews (68 mothers and 99 fathers), 58 endline surveys with fathers, and 84 in-depth interviews (37 midline, 47 endline) with mothers, fathers, and group leaders. Collectively, these sources encompass both quantitative and qualitative data and provide a broad assessment of program implementation across multiple time points during and after the intervention and from various perspectives, including fathers, mothers, facilitators, and M&E officers.

**Table 1.** Overview of data collected as part of the process evaluation.

| Data source | Respondent | Frequency or time frame | Number of records | Questions included to assess process evaluation dimension(s) |  |  |
| --- | --- | --- | --- | --- | --- | --- |
|  |  |  |  | Feasibility | Acceptability | Appropriateness |
| **Attendance form** | Facilitator | Each parenting group session | 6 groups x 15 sessions = 90 records | Presence/absence of each participant<br>Reason for participant absence (if applicable) |  |  |
| **Participant dropout form** | Facilitator | At the time of participant dropout (as cases arise) | 2 couples | Name of participant wishing to dropout<br>Reasons or context surrounding why this participant is dropping out from the program |  |  |
| **M&E observation form** | M&E officer | Each parenting group sessions (when M&E officer is present) | 75 observations across 6 communities<br>(13 in Village 1, 13 in Village 2, 14 in Village 3, 9 in Village 4, 15 in Village 5, 11 in Village 6,) | Caregiver engagement levels during the session<br>Caregiver enjoyment during the session<br>Any modifications made to the session<br>Adherence to the session guide<br>Handouts distributed to caregivers<br>Homework assigned to caregivers | Facilitator effectiveness in encouraging group participation<br>Facilitator effectiveness in using visual aids during the session |  |
| **Facilitator feedback form** | Facilitator | After each parenting group session | 79 feedback forms across 6 communities<br>(13 in Village 1, 14 in Village 2, 13 in Village 3, 14 in Village 4, 13 in Village 5, 12 in Village 6) | Adherence to the session guide<br>Any modifications made to the session |  |  |
| **Participant feedback form (i.e., exit interviews)** | Subset of participating mothers and fathers | After each parenting group session | 167 exit interviews<br>(68 with mothers and 99 with fathers) | Handouts received by caregivers<br>Contact with the group leader in the past week | Session rating<br>Facilitator's ability to clearly explain the session activities and content |  |
|  |  |  |  |  | Facilitator's readiness and confidence in delivering the session<br>Facilitator effectiveness in using images and other materials during the session |  |
| **Endline quantitative survey** | Enrolled fathers | At endline | 58 fathers | Frequency of contact with the group leader<br>People who remind fathers about the session time/date | Program rating<br>Satisfaction with the session duration<br>Satisfaction with the session frequency (weekly sessions)<br>Influence of incentives on session attendance | Preferred session format (mixed-gender sessions vs same-gender sessions)<br>Comfort with female vs male facilitator<br>Sharing of session information<br>Individuals with whom fathers shared session information<br>Topics discussed with others<br>Topics perceived as misaligned with fathers' values or beliefs<br>Fathers' favorite topics |
| **Qualitative in-depth interviews** | Subset of enrolled mothers and fathers | At midline and endline | Midline: 37 total interviews<br>(6 group leaders, 12 high attending fathers, 6 high attending mothers, 13 low-attending fathers)<br>Endline: 47 total interviews<br>(14 baseline fathers, 6 star fathers, 15 baseline mothers, 6 star mothers, 6 group leaders) | Relationship with the group leader<br>Group leader's responsibilities<br>Facilitator's responsibilities | Program satisfaction<br>Caregivers' experience in the program<br>Facilitator's effectiveness in leading the sessions<br>Suggestions for improving facilitator's session delivery<br>Perceived ease of attending the sessions<br>Facilitators and barriers to attendance | Acceptability of the session topics<br>Satisfaction with the materials used in the sessions<br>Use of handouts after sessions<br>Comfortability with mixed-gender sessions |

Quantitative process data were collected at each session and at endline. At each session, the facilitators recorded attendance. At the end of each session, facilitators, participants and the M&E officer completed tablet-based surveys that were programmed on KoboToolbox. Facilitators completed a session feedback form that captured session quality, ease of delivery, recommended revisions to the guide and overall impressions. Throughout each session, the M&E officer observed and documented the performance of the facilitator, participant engagement, adherence to the session guide and overall session quality. At the end of the session, the M&E officer randomly sampled about three caregivers, rotating selections to avoid repeated interviews, to complete a brief (∼10 minutes) survey capturing their overall satisfaction and feedback. In total, at the end of each session, process data were systematically collected using four forms: (1) attendance form, (2) facilitator feedback form, (3) M&E observation form, and (4) participant feedback form. At endline, all enrolled fathers were interviewed to assess overall program satisfaction, content acceptability, and satisfaction with the program model.

Qualitative process data were collected at midline and endline. Two independent Tanzanian research assistants interviewed caregivers at midline to capture their overall experience in the program and their feedback on various components of the program, i.e. the videos, handouts, toys, images, etc. To comprehensively capture diverse perspectives, we sampled caregivers with high and low attendance. From each community, 6 caregivers were selected: 2 fathers with low attendance, 2 fathers with high attendance, 1 mother with high attendance and 1 group leader. Low attendance was defined as attending fewer than half of the sessions by the time of the interview, while high attendance was defined as attending more than half. At endline, we followed up with caregivers who were randomly sampled at baseline and additionally included 2 “star caregivers” (one mother and one father) from each community. “Star caregivers” were identified by facilitators as individuals with high attendance and consistent engagement in the program.

Both midline and endline in-depth interviews were conducted in Kiswahili and lasted approximately one hour. These interviews were audio recorded using handheld voice recorders and later translated directly into English by experienced Tanzanian research assistants and TAHEA-Mwanza staff. These transcripts were further reviewed by a member of our team who is fluent in both languages and provided contextual insights and further revised the translations throughout the analysis to ensure accuracy and depth.

### Data analysis

Our analysis was organized around the three focal dimensions of process evaluation: feasibility, acceptability, and appropriateness. For each dimension, we triangulated data across multiple sources and compared findings between fathers and mothers to explore any potential gender differences. By drawing on similar or complementary questions across tools and respondent types (e.g., participants, facilitators, and M&E officers), we assessed each dimension from multiple perspectives to identify converging or diverging patterns in reported experiences.

#### Quantitative analysis

Quantitative process data were analyzed using R. Descriptive analyses were conducted to summarize caregiver responses, presenting proportions overall, as well as disaggregated by gender for each relevant variable to compare perceptions and experiences between fathers and mothers. Additionally, open-text responses in the feedback forms (i.e., reasons for absence) were categorized into thematic groups to organize caregiver responses. This categorization enabled us to tabulate and quantify the number of caregivers reporting similar responses.

#### Qualitative analysis

Qualitative data were analyzed using ATLAS.ti through an immersive and iterative team-based approach. Following midline data collection, a team of six analysts collaboratively developed a codebook, which was iteratively refined after each researcher independently coded several transcripts. Weekly meetings were held to resolve coding questions, revise definitions, and add new codes as needed. Two researchers reviewed all coded transcripts to address any discrepancies flagged by the analysts and to ensure consistent code application. A similar process was followed for the endline interviews involving four analysts. Both the midline and endline interviews were double coded independently by two researchers to ensure consistency and reliability.

Following coding of both midline and endline interviews, we conducted an in-depth thematic content analysis. Detailed summaries were prepared for each code relevant to the process evaluation to identify common themes. Findings were disaggregated by gender to compare perspectives between mothers and fathers and any potential gender differences that may have emerged. Throughout the analysis process, both US-based and Tanzanian team members collaborated closely to ensure contextual relevance and enhance interpretive rigor.

### Ethical approval

This study received ethical approvals from the Institutional Review Board at Emory University and the National Health Research Ethics Committee at the National Institute of Medical Research, Tanzania. Written informed consent was obtained from all study participants.

## RESULTS

### Demographic characteristics

Baseline demographic characteristics of the 61 enrolled couples and their children are summarized in **Table 2**. The median age of the index children was 12 months, and a greater proportion were girls (54%) versus boys (46%). The median age of the fathers and mothers was 37 years and 28 years, respectively. Parental education was relatively low, with only 15% of fathers and 12% of mothers having fully completed secondary school. Regarding couples’ relationship status, most caregivers were partnered and cohabitating but not formally married (84%), while 16% were legally or formally married. The median duration across all partnered relationships was 7 years. In addition to the enrolled index child, the majority of caregivers (87%) had another child, with a median household size of three children.

**Table 2.** Demographic characteristics of participants at baseline as reported by fathers.

| **Sociodemographic characteristics** | **N (%) or median [IQR]** |
| --- | --- |
| Father characteristics |  |
| Father's age (in years) | 37 [28, 43] |
| Father's highest level of education |  |
| No education | 2 (3.3%) |
| Some primary school | 9 (14.8%) |
| Primary school completed (or some secondary) | 41 (67.2%) |
| Secondary school completed or higher | 9 (14.8%) |
| Mother characteristics |  |
| Mother's age (in years) | 28 [24, 34] |
| Mother's highest level of education |  |
| No education | 4 (6.6%) |
| Some primary school | 6 (9.8%) |
| Primary school completed (or some secondary) | 44 (72.1%) |
| Secondary school completed | 7 (11.5%) |
| Child characteristics |  |
| Child's age (in months) | 12 [7, 18] |
| Child's gender |  |
| Female | 33 (54.1%) |
| Male | 28 (45.9%) |
| Relationship/household characteristics |  |
| Marital status |  |
| Legally or formally married | 10 (16.4%) |
| Not legally or formally married but living with<br>a partner in a consensual union | 51 (83.6%) |
| Length of marriage/co-habitation (in years) | 7 [3, 14] |
| Number of children in the household | 3 [2, 5] |

### Feasibility

Based on the facilitator feedback and M&E observation form, the majority of the program was delivered as intended. Fathers and mothers reported that the facilitators and group leaders fulfilled their roles in line with the program model. Session attendance was acceptable, with fathers attending an average of 9 out of the 15 sessions and mothers attending 9 out of 13 sessions. The main barrier to fathers’ attendance was work-related conflicts.

#### Overall session attendance and engagement

The program consisted of 15 total sessions, 13 designed for couples and 2 for fathers only. The average attendance rate was slightly higher for mothers (65.8%) than fathers (60.7%) (**Figure 1**).

**Figure 1.**
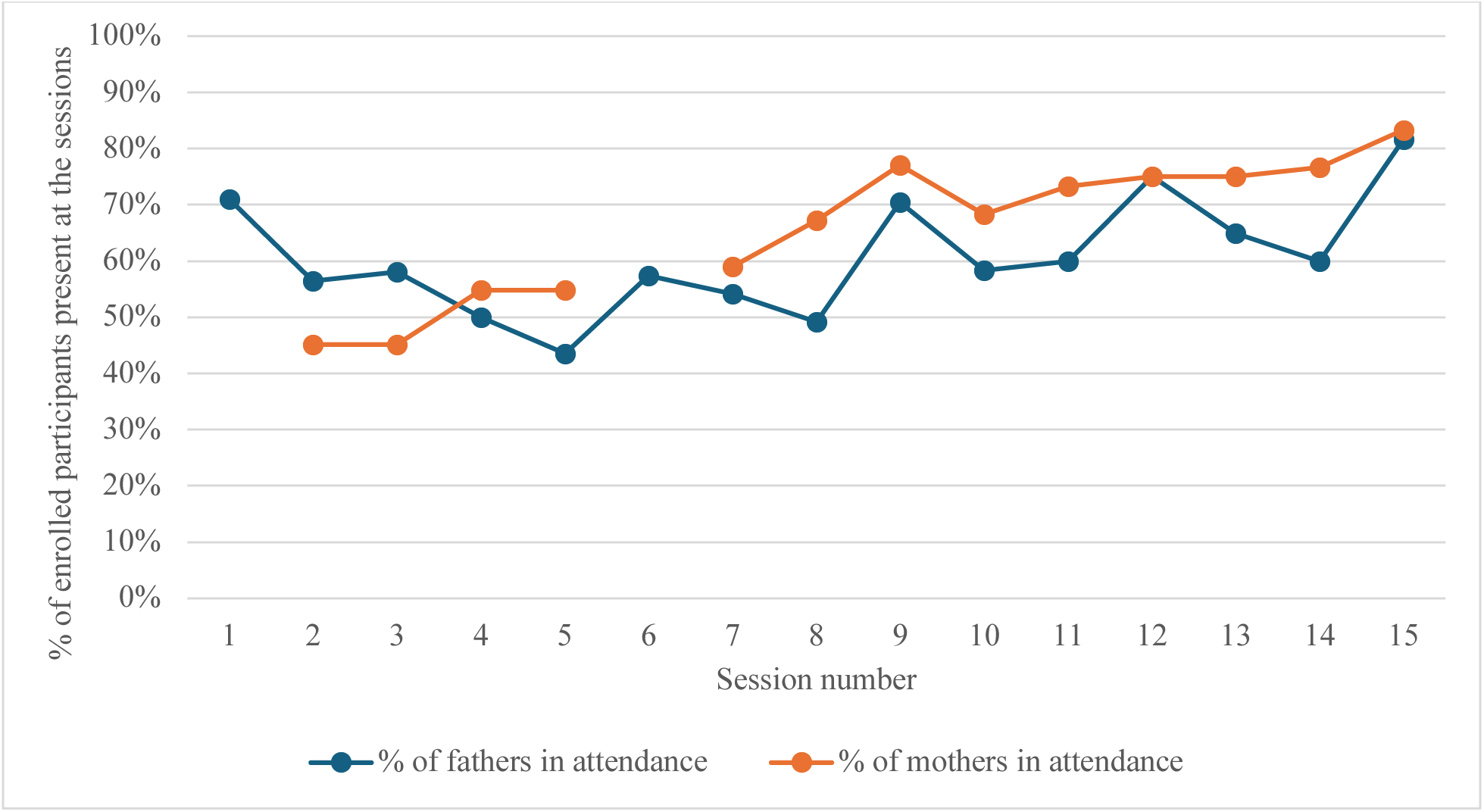
Average attendance rates for fathers versus mothers across sessions. *Note: Data on mothers’ attendance are unavailable for sessions 1 and 6, as these were fathers-only sessions and are therefore not shown*.

Based on facilitator’s reports on attendance forms, the most common reason for fathers’ absence was work or other related activities (44%), followed by being unreachable (17%), and travel (15%) (**Figure 2**).

**Figure 2.**
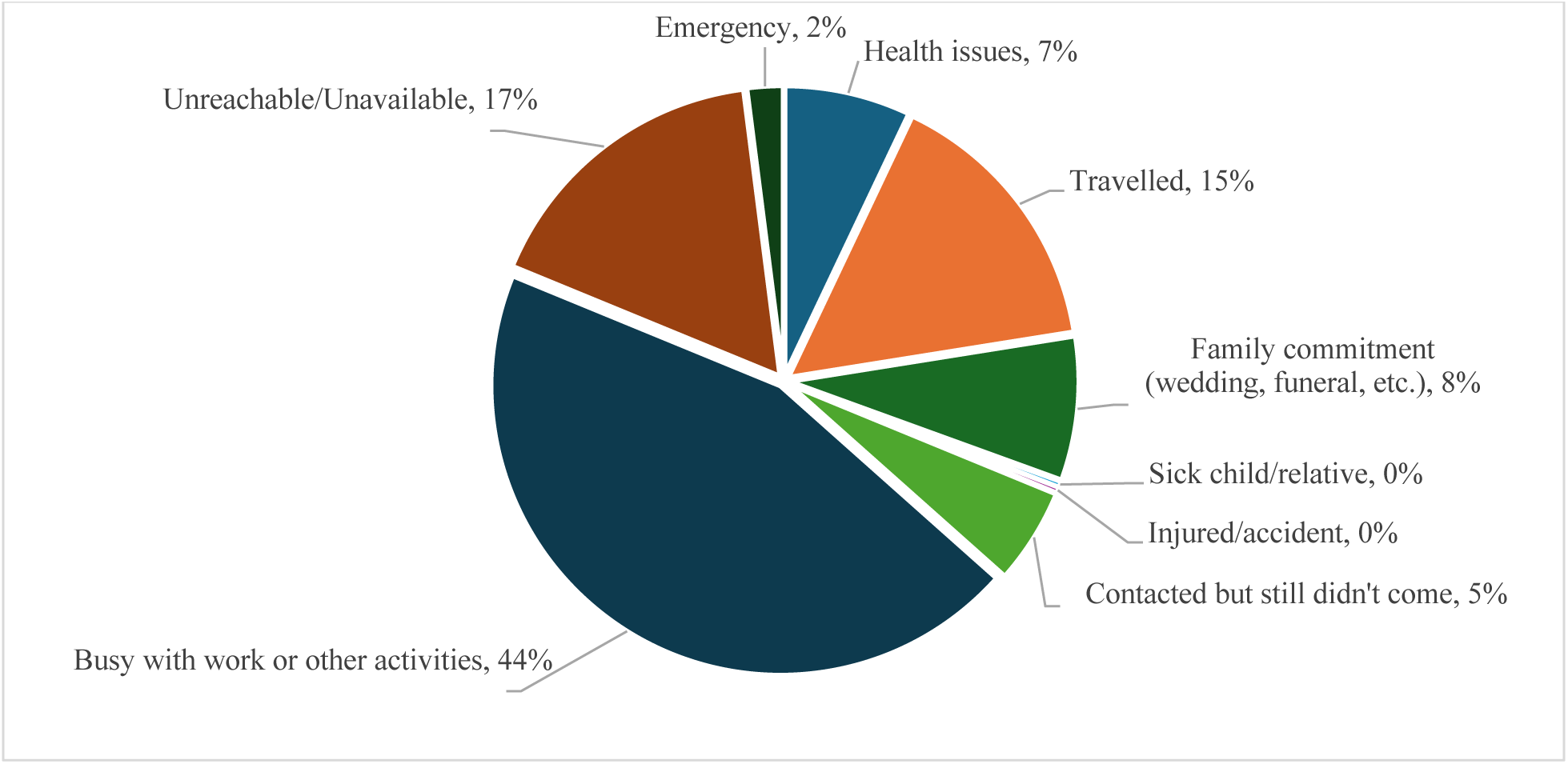
Reasons for fathers’ absence during the program, based on attendance records of those absent at any session over the duration of the program (n=299).

These findings were also reflected in the midline and endline qualitative interviews, where most fathers reported work-related schedule conflicts as the primary barrier to attendance.

> *“As a father, I am the breadwinner in the family and our community views me as such, therefore sometimes I may still be engaged [with work] while the program is in session. So, that may limit my attendance.”* (Father #7 at midline, Village #5)

Regarding mothers’ absence – based on the exit surveys and midline and endline qualitative interviews – the most commonly reasons were household chores and caregiving responsibilities, family emergencies, caring for a sick relative or child, and attending funerals. For example, one mothers shared, *“Searching for water, doing housework and raising children takes a lot of time. I can even be busy during the time for the session.”* (Exit interview, woman in Village #5)

Despite the moderate participation described above across all 15 sessions, the M&E feedback forms indicated that 86.3% of the sessions had good or excellent engagement among both fathers and mothers. Additionally, in approximately 94.6% of sessions, the M&E officer rated that the majority of fathers appeared to enjoy the session. Similarly, in about 94.8% of the sessions, the majority of mothers were rated as enjoying the session.

#### Retention

Of the 62 couples originally enrolled in the intervention, there were two dropout cases, resulting in a retention rate of 96.8%. In one case, a father was asked to leave by the village chairperson and fellow group members because he was deemed “unfit” for the group, and his partner was also withdrawn from the program. In the other case, a father no longer wished to participate in the program and dropped out at midline. Since neither he nor his partner had attended any session, the couple was subsequently withdrawn from the program.

### Fidelity

#### Session structure and delivery

According to the facilitator feedback and M&E observation form, 85% and 73% of all sessions, respectively, were delivered exactly as outlined in the session guide (**Table 3**). The remaining sessions were delivered with more than half of their content adhering to the guide. Any changes made to the guide were relatively minor and included small modifications to improve clarity, add discussion questions, introduce energizers between activities, and to provide additional examples and opportunities for caregivers to role play. *“After activity 2 the facilitator introduced a short break game whereby the participants had to catch a ball after he saw that they were not fully participating. This improvisation brought back the participants’ attention in the class, and they started responding positively.”* [From the M&E observation form] In a few instances, facilitators skipped a role play activity due to time constraints.

**Table 3.**
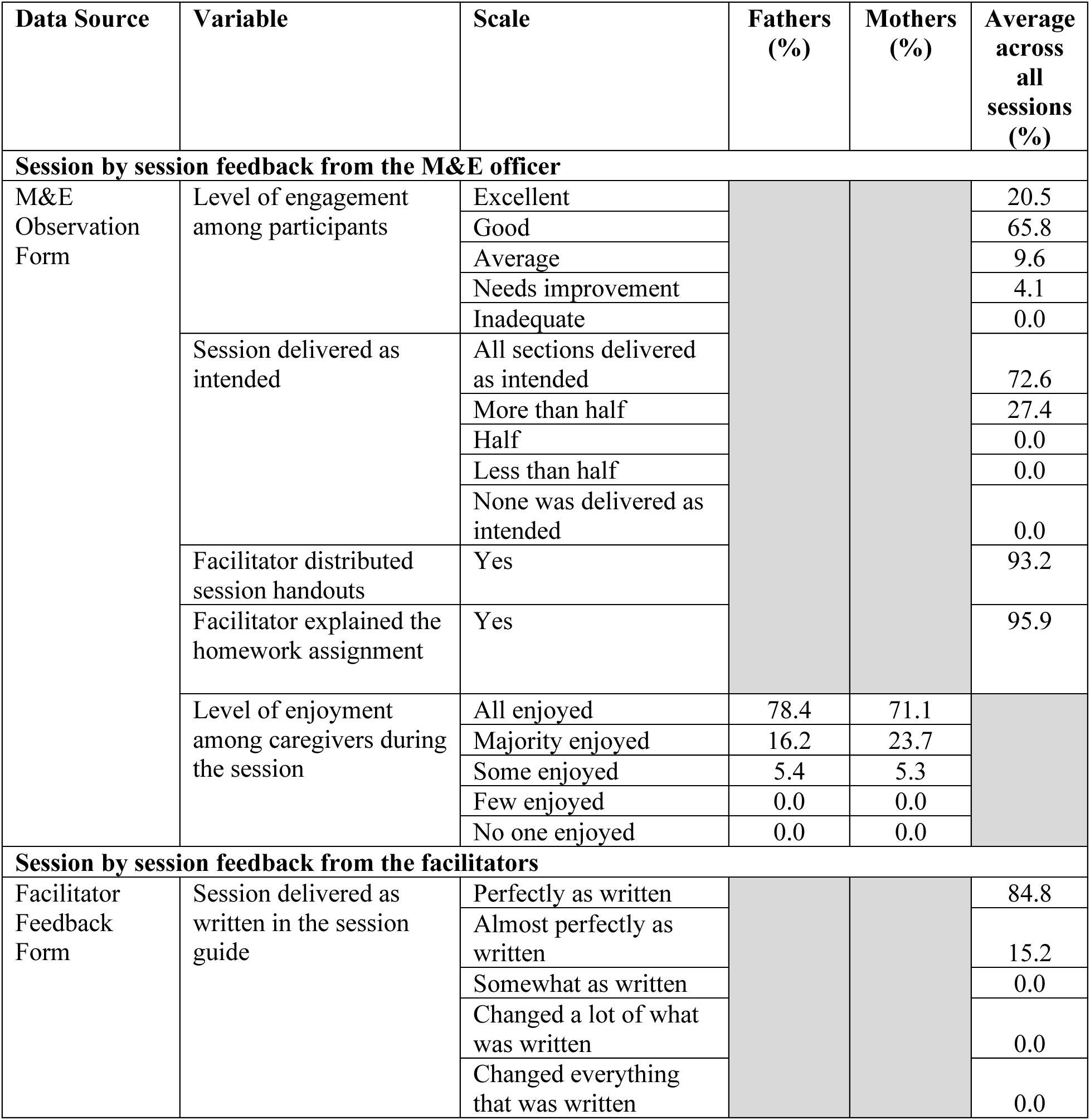
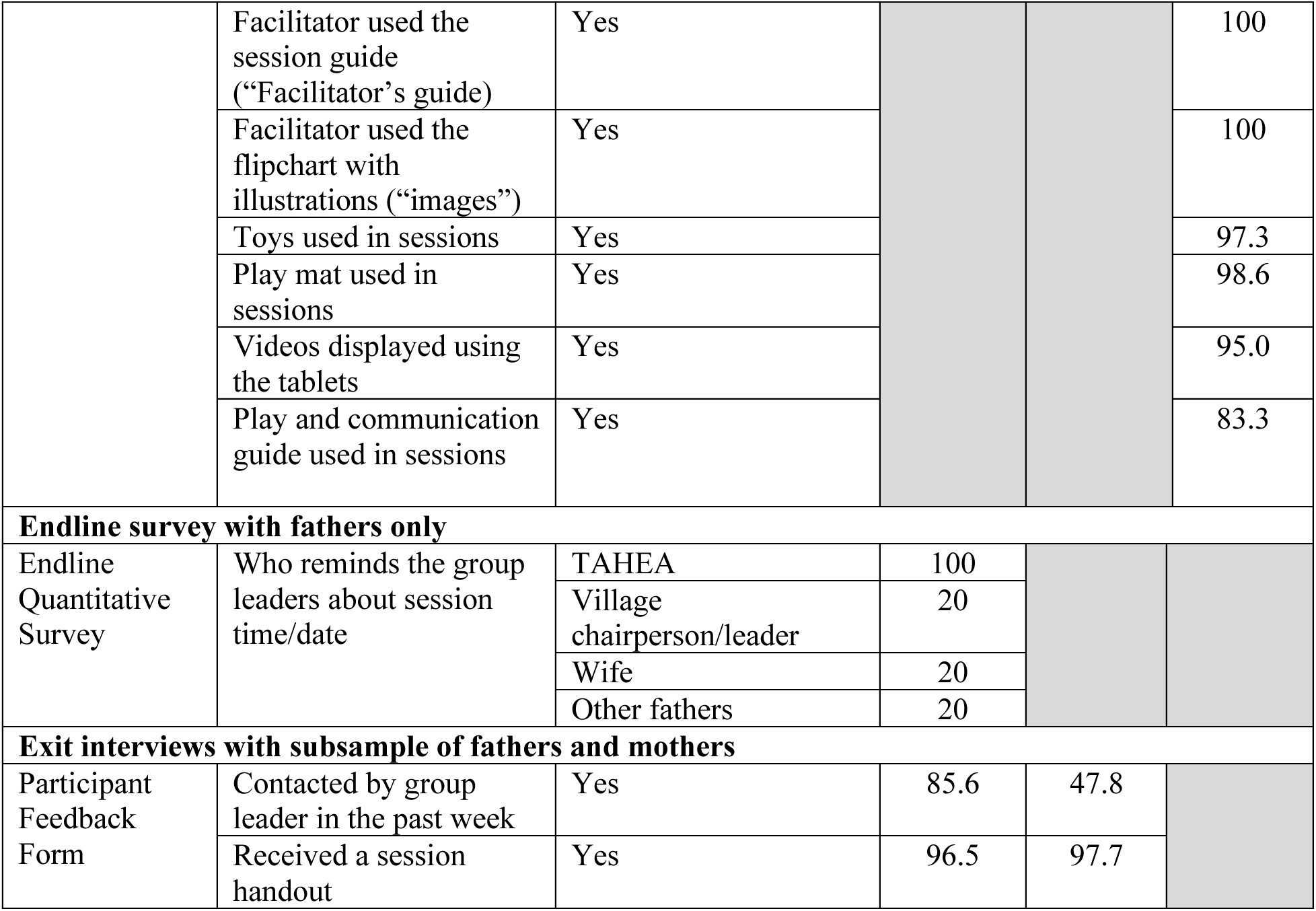
Session fidelity, Participant engagement, and Fulfillment of roles by group leaders and facilitators.

#### Program materials

According to the facilitator feedback form, both the session guide and flipchart with illustrations were used in every session. The toys and play mat were used in approximately 97.3% and 98.6% of couple-based sessions, respectively. Additionally, as outlined in the curriculum, 5 out of the 6 (83.3%) intervention communities used the Play and Communication guides as part of session 8 (**Table 3**). In the one community where the guides were not used, the play activity was skipped due to a limited number of children present during the session. In contrast, the videos were shown in 95% of the sessions where they were included as part of the curriculum, demonstrating high engagement with this resource throughout the program.

In addition to the session materials, as reported in the M&E observation form, the facilitator explained the homework assignment and distributed the session handout in approximately 95.9% and 93.2% of all sessions, respectively. This was corroborated by caregivers, with 96.5% of fathers and 97.7% of mothers who completed the exit interviews confirming that they received a handout at the previous session.

#### Messages received

When asked what they learned during the program in the midline and endline qualitative interviews, caregivers recalled messages that closely reflected the core content of the curriculum. Both fathers and mothers consistently described lessons spanning the full range of session topics, including ECD, gender roles, couples’ relationships, parent-child relationships, mental health, nutrition, and budgeting. This suggests that program content was delivered as intended and well understood by participants.

The majority of caregivers described learning about ECD, the importance of building strong relationships with their children, and specific activities to support early child learning. For example, fathers and mothers recalled stimulation activities such as making homemade toys, engaging in play, and some also mentioned the importance of responsive caregiving interactions such as noticing and responding to children’s cues.

> *“I have learnt a lot, for example making playing toys like a bell for my child, also the issue of training the baby on understanding things by their names. […] For instance, if you give a toy to your baby and he or she refuses to take and she needs a ball, then you have to follow what she needs and give it to him or her, even when the baby learns to walk or crawl it is good to congratulate him or her to make him or her motivated and improve.”* (Father #7 at endline, Village #4)

When describing lessons on responsive caregiving, multiple caregivers specifically referred to the videos as helping to depict and clarify this concept for them.

> *"There’s one video I remember. It was about a man named John. They showed us a video of him playing soccer, and he was playing with the intention of cheering up the child. But the child, instead of continuing with the ball, he wanted to go to a nearby trolley. He [the child] left his ball and started playing with that trolley. The lesson we learned from that was to follow what the child wants in order to make them happy. So, the man had to leave his ball and start pushing the trolley with the child. We were taught many things in that video."* (Father #1 at midline, Village #4)

Messages about gender roles and shared responsibilities were also salient, particularly among fathers. A common message recalled was learning about the importance of men being more actively involved in caregiving and domestic tasks, not only as financial providers, but also as emotionally present fathers who spend quality time with their children and families.

> *“Taking an example of session 8, we saw a picture that was about a certain father who was a motorbike driver who had no time with spending some moments with his family, he was just giving money for family expenses and leave without even playing with his children or spending some time with them. […] On the other hand, we saw a picture also concerning another motorbike driver who was caring for his family, he used to set time for spending some moments with his children and the whole family and the family loved him very much.”* (Father #9 at midline, Village #2)

Fathers and mothers also frequently described lessons learned on positive couples’ relationships, including communicating respectfully with their partners to resolve conflicts and joint decision-making, particularly around finances, such as budgeting and saving. Several caregivers also expressed a deeper understanding of the harmful effects of intimate partner violence. Mental health was another key theme, especially among fathers, who recalled learning various stress management and coping strategies (e.g., seeking advice from trusted friends, taking personal time to relax) and the risks of negative coping strategies such as heavy drinking. Importantly, these mental health lessons were often discussed in the context of relationship stressors, with participants describing that the program taught them strategies to better navigate emotional challenges (i.e. misunderstandings, conflicts) within their marriages.

> *“We were also taught on how to handle our misunderstandings, when you have misunderstanding with your partner it is good to sit down with her and solve or go to your neighbor whom you trust and share with him or her your family misunderstandings. That way it can help you can get advice from him or her and help yourself to vent and get a relief. Sometimes one may not be having any friend and you find his wife has confused him, so you have to go out of home premises for some walks so that you can have a relief.”* (Father #7 at endline, Village #4)

Many caregivers also reported gaining knowledge about child nutrition and more specifically ensuring balanced diets for young children and exclusive breastfeeding. For example, one father shared, *“We learnt that a young baby from zero to six months of age should be breast fed and as he/ she grows above she or he has to be given food comprising various food categories like vegetables, fruits, protein and carbohydrate.”* (Father #9 at midline, Village #2)

While the topics overall were delivered with fidelity, some areas like responsive caregiving and discipline were recalled less consistently or with less detail relative to their emphasis in the curriculum. Caregivers often described positive parenting in terms of engagement in play and communication, but many were less explicit about the quality of these parent-child interactions, particularly the importance of responding to and following the child’s lead. One father, however, well-articulated what he had learned about following his child’s cues and supporting learning in everyday activities,

> *“[I have learned] for instance if the baby is pointing at a tree you should mention to him/ her the name tree, if he or she is pointing at a bird you should mention bird so that he/ she becomes familiar with the names of such things/ objects. […] You have to follow what she needs and give it to him or her, even when the baby learns to walk or crawl it is good to congratulate him or her to make him or her motivated and improve.”* (Father #7 at baseline, Village #4)

#### Group leaders’ roles in the program

Through the midline and endline qualitative interviews, most fathers and mothers clearly articulated the role of the group leader, providing evidence that group leaders fulfilled their expected responsibilities. For example, participants like the following farther explained that group leaders reminded participants of session times and days:

> *“He [the group leader] had a responsibility of informing us what was happening about the meeting plans as a coordination between us as group members and TAHEA, and whenever one gets an emergency of not attending any meeting once you inform. He understands and normally reports to TAHEA people on the excuse given by the absentee.”* (Father #6 at endline, Village #4)

Many caregivers and group leaders stated that these reminders were communicated via SMS, phone calls and sometimes home visits.

> *“Our group leader used to call us on phone call or sometimes he comes direct at home or sometimes I also make a call to him to ask about any group session timetable.”* (Mother #4 at midline, Village 2)

However, group leader roles were carried out differently for and with greater attention to fathers than mothers. Among exit interview respondents, 85.6% of fathers reported being contacted by a group leader in the past week, compared to only 47.8% of mothers (**Table 3**).

Qualitative interviews at midline and endline with mothers further supported this conclusion, with some explicitly stating that the group leaders contacted their male partners instead of them directly.

> *“He [the group leader] would send the messages to my husband’s phone only. I would get the information through my husband, who would inform me about the sessions.”* (Mother #7 at endline, Village #3)

While group leaders reported no difference in ease of communication between fathers and mothers, they noted contacting fathers more often, usually because they only had fathers’ phone numbers and found it more convenient.

> *"I do not normally call women on phone. I usually call men because I do not have women’s contact numbers."* (Group leader at midline, Village #3)
>
> *“To be honest, I do not have much time to talk with the women. I rarely communicate with them, mostly when we gather for sessions, which is when I try to talk to them.”* (Group leader at midline, Village #1)

### Acceptability

Based on the endline quantitative survey, all fathers rated the program favorably, with 90% rating it as “very good” and 10% as “good” (**Table 4**). In midline and endline qualitative interviews, both fathers and mothers expressed appreciation for the program content, facilitators, and group leaders. However, a primary source of dissatisfaction among caregivers, that largely emerged from the midline qualitative interviews, was the lack of incentives during the first half of the program, which persisted at endline in communities that did not receive incentives.

**Table 4.**
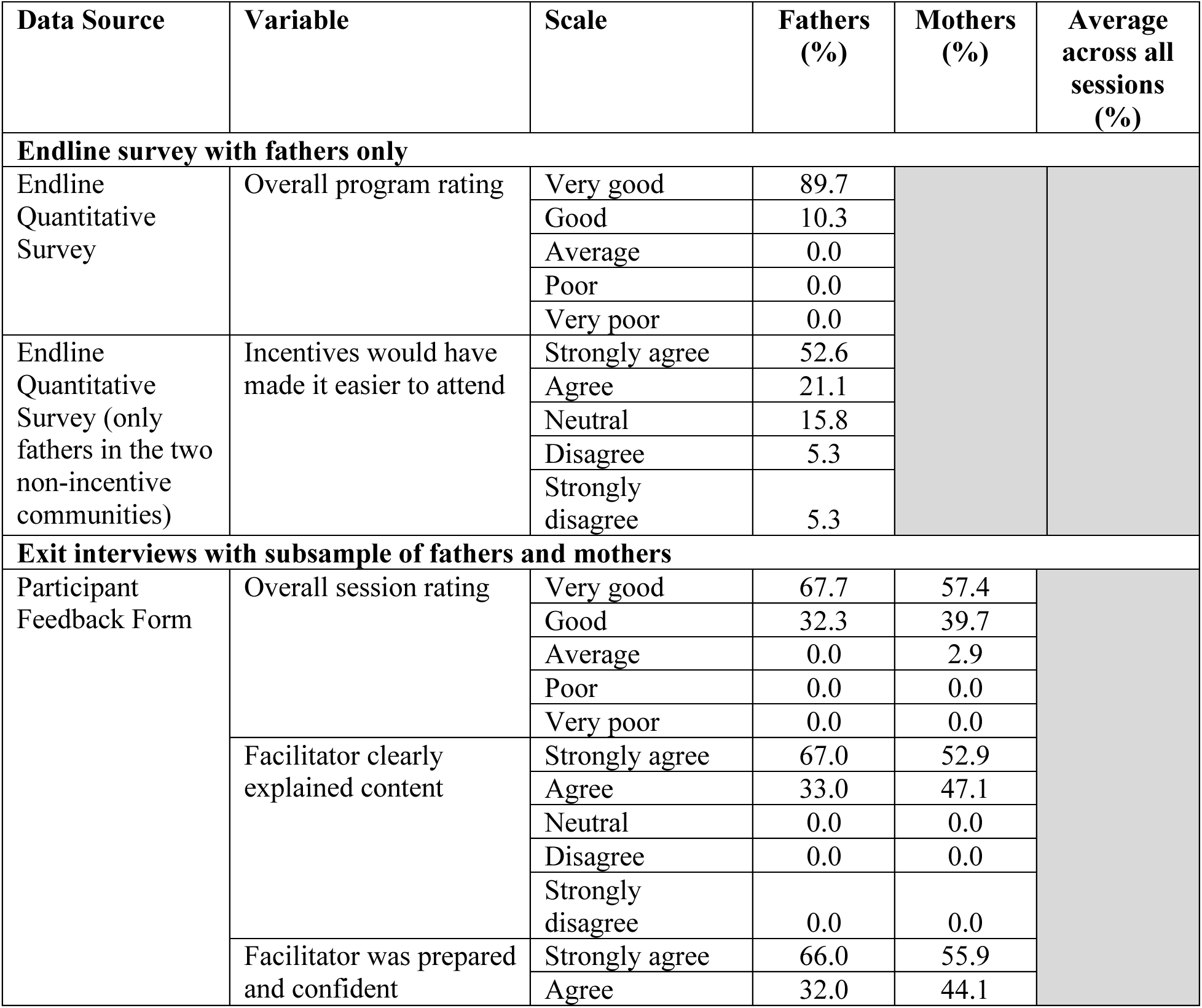
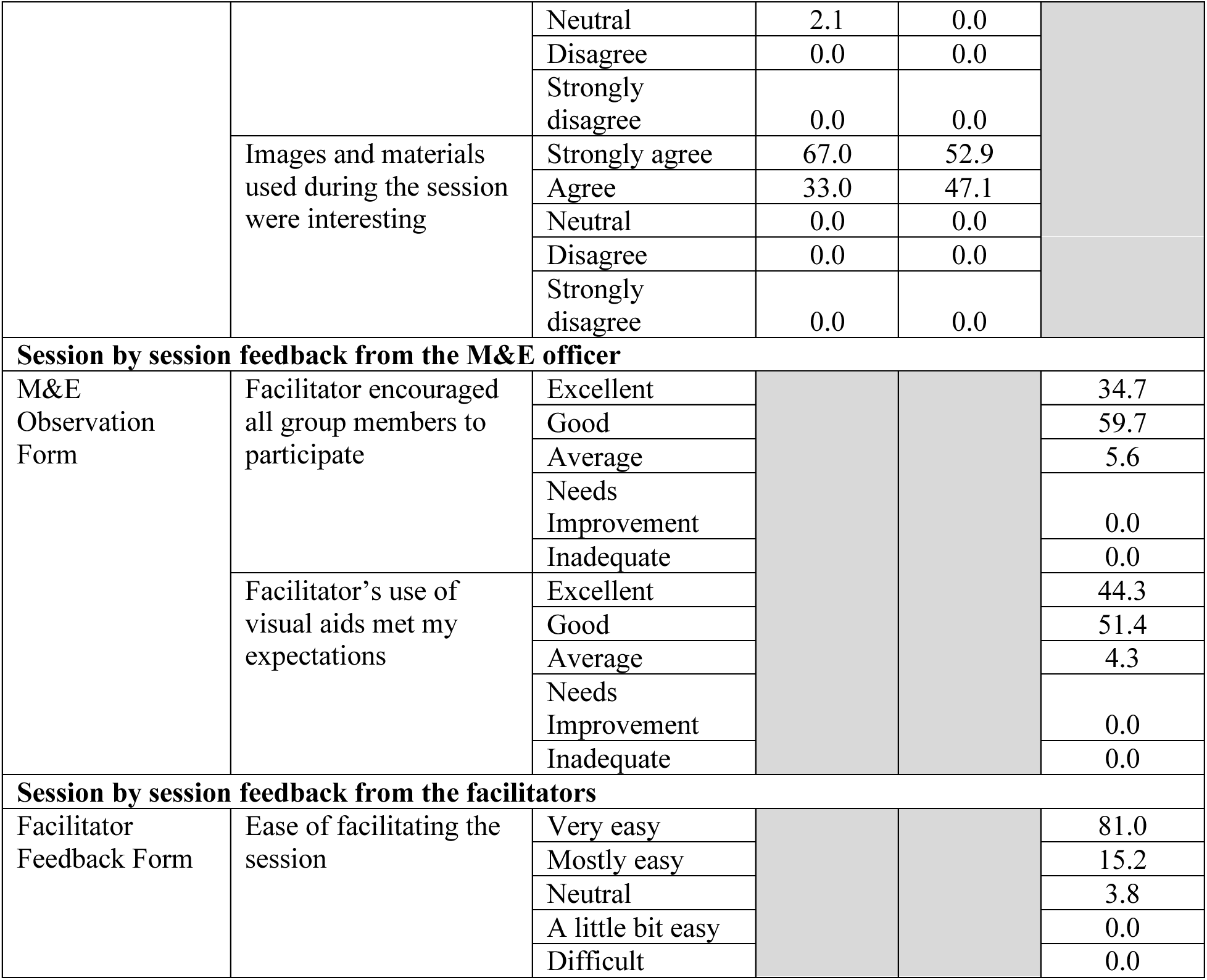
Satisfaction with the program & materials, quality of facilitation, and ease of session delivery.

#### Group session facilitation

Based on the exit interviews, all fathers and mothers agreed that the facilitators clearly explained the session activities and content. Additionally, all mothers and 98% of fathers agreed or strongly agreed that the facilitators were well-prepared and confident in delivering the sessions (**Table 4**). Midline and endline qualitative interviews further highlighted caregivers’ appreciation for facilitators’ use of practical activities (e.g., role plays, playing with the children), training materials (e.g., toys, videos, flipcharts), community-relevant examples, review of key takeaways from the previous session, and integration of earlier concepts into current sessions.

For example, one father described how the facilitator consistently reviewed previous content to ensure understanding before moving on:

> *“Mr. X [the facilitator] I would rate him at 100%. This is because whenever he came to a session he never moved on with any topic without doing a recap or review of the previous session, so before you continue with another topic you spend like fifteen minutes doing revision of the previous topic and clear all issues that were not clear and then move on with the new topic, so that is how it was.”* (Father #11 at endline, Village #4)

Similarly, one mother a mother emphasized the facilitators’ use of practical, visual, and hands-on activities to enhance understanding and engagement:

> *“I give her all 100%. She was teaching very well, nothing was left, she was teaching them all, giving examples and showing pictures, it was easy for us to understand, even if someone did not understand by words, but by seeing pictures you understand We were even drawing pictures, she brought piece of clothes for us to make dolls, she taught us the way to make impressing playing tools.”* (Mother #7 at endline, Village #6)

In addition to their facilitation skills, both fathers and mothers shared, in the endline qualitative interviews, that the facilitators were friendly, approachable and patient with the participants. Thus, providing them with sufficient time to ask questions and discuss the content during the sessions. For example, one father shared:

> *“They had a big and tough role, they did it to their level best, fortunately they succeeded in high percentage. Because managing explaining something to someone who you are not living together and the person understand you, it is not an easy work, sometimes someone come with stress, but they managed to make us feel ok, to change our thoughts, they worked hard. Because they were teaching well, charming and active…”* (Father #1 at endline, Village #5)

Another mother highlighted how her facilitator was respectful and created a supportive atmosphere during sessions.

> *“I was happy with facilitators, they were not harsh, they were teaching as if they know us, also participants were not arguing, we were respecting everyone’s opinion, we respected each other, we were close to each other, that is the strength I can say . . . Everyone was asking or responding freely and was respected.”* (Mother #7 at endline, Village #1)

M&E feedback forms further corroborated these findings, showing that facilitators effectively encouraged all group members to participate in 94% of sessions and used visual aids effectively with ratings of “good” or “excellent” in 96% of the sessions (**Table 4**). Fathers and mothers also reported that session materials were engaging, with all fathers and 97% of mothers rating sessions as “very good” or “good.” Facilitators self-reported that 96% of sessions were easy to facilitate.

Qualitatively, a few fathers at endline however a minor dissatisfaction with the group facilitators, primarily due to the late arrival by the facilitators. For instance, one father shared, *“I didn’t give them this 5% [of satisfaction] because sometimes they were coming late, participants were coming on time, but the facilitators were coming late… Yeah, if not two sessions, then is three sessions they came late, so that 5% is due to time management, no other issues”* (Father #1 at endline, Village #5) However, other caregivers noted that facilitators were generally punctual, such as this example from another father, *“Honestly, I was very impressed with their teaching. They taught us very well, provided excellent lessons, and worked hard to help us. They were always on time. We would arrive late, but they were always on time, and we were happy with what we learned.”* (Father #7 at endline, Village #6)

#### Provision of incentives

One key area of dissatisfaction was the lack of incentives especially during the first half of the program. Both mothers and fathers, regardless of their attendance levels, suggested that incentives, loans or capital could be offered to help boost attendance among fathers and mothers.

> *“Men could just look at the burdens of the family, because in most of the households, men are the income earners, they go out in the morning searching for income to make sure the family doesn’t starve. And we requested that due to the challenges occurring with attendance, if I attend the group, at least I should be given something to support me. At least if I spend my day attending the sessions, I should be given some amount of money to cover the gap for the income I would have earned to support my family.”* (Father #7 at endline, Village #3)
>
> *“You may find that someone postponed her chores just to attend the session, that is why I have told you that, some people were not attending, not that they intend, but because of poor income. She will decide to miss classes because she needs to work for her family.”* (Mother #7 at endline, Village #6)

This was especially true in the two non-incentives communities, where 74% of fathers in the endline quantitative survey agreed that incentives would have made it easier for them to attend the sessions (**Table 4**).

Qualitatively at endline, some fathers and mothers explained that their expectations for financial incentives were set by prior experiences with similar programs that offered such support.

> *“Many people have participated in projects like X [project name], and when they were invited to seminars, they would even get 5,000 or 10,000 shillings if the seminar was from morning to evening. (…) That’s why they were used to it. So, when they hear that a certain organization is coming, they only think about the money. They believe they will always get something.”* (Mother #6 at endline, Village #1)

### Appropriateness

The majority of fathers and mothers found the session content and program model to be appropriate, aligned with their parenting needs, and contextually relevant. They reported sharing the session information with others including those outside of the program. In addition to the diverse topics, caregivers appreciated the various sessions materials because they clarified key messages, reflected their experiences, and offered practical ways to apply lessons in daily life.

A distinct feature of this program was its couples-based approach, with a specific focus on engaging fathers. Thus, most sessions were delivered in mixed-gender sessions by either a female or male facilitator. The majority of caregivers reported feeling comfortable participating with their partners. There was no strong preference for either a male or female facilitator, with most caregivers being comfortable learning from both male and female facilitators.

#### Session content

The session content was highly acceptable to most fathers and mothers. Qualitatively at endline, many caregivers described all program topics as relevant and important to their lives. Additionally, they reported that those with whom they shared the program messages responded positively and expressed interest in joining the program.

> *“They [the session topics] are accepted. For instance, the topics I mentioned, like violence and budgeting, are very relevant to the community. People encounter these issues often. After learning, we educated others, and some have changed.”* (Mother #11 at endline, Village #3)

In the endline quantitative survey, fathers most frequently ranked responsive caregiving, economic strengthening, mental health, intimate partner violence, and fatherhood as their most interesting topics learned (**Table 5**). Qualitatively at endline, many fathers particularly valued the session on economic strengthening for its practical guidance on managing household finances, especially balancing money for saving vs spending.

**Table 5.** Top-rated session topics, sharing of session content and perceived appropriateness and satisfaction with the program structure/format.

| Data source | Variable | Scale | Fathers (%) | Mothers (%) |
| --- | --- | --- | --- | --- |
| **Exit interviews with subsample of fathers and mothers** |  |  |  |  |
| Participant Feedback Form | Preferred format for group sessions | Mixed-gender sessions | 90.0 | 92.0 |
|  |  | Same-gender sessions | 10.0 | 8.0 |
|  | Discussed session content with others | Yes | 71.0% | 77.8% |
| **Endline survey with fathers only** |  |  |  |  |
| Endline Quantitative Survey | Satisfaction with session frequency (once per week) | Very satisfied | 32.8 |  |
|  |  | Satisfied | 58.6 |  |
|  |  | Neutral | 0.0 |  |
|  |  | Dissatisfied | 8.6 |  |
|  |  | Very dissatisfied | 0.0 |  |
|  | Satisfaction with the 2-hour session duration | Very satisfied | 32.8 |  |
|  |  | Satisfied | 65.5 |  |
|  |  | Neutral | 0.0 |  |
|  |  | Dissatisfied | 1.7 |  |
|  |  | Very dissatisfied | 0.0 |  |
|  | Discussed session content with others | Yes | 96.5 |  |
|  | People fathers shared session content with | Friends | 70.9 |  |
|  |  | Neighbors | 61.8 |  |
|  |  | Partners | 27.3 |  |
|  | Session topics most repeatedly ranked among fathers' top three favorites | Responsive caregiving | 51.7 |  |
|  |  | Economic strengthening | 43.1 |  |
|  |  | Mental health | 36.2 |  |
|  |  | Intimate partner violence | 25.9 |  |
|  |  | Fatherhood | 19.0 |  |
|  |  | ECD | 8.6 |  |
|  |  | Discipline | 5.2 |  |
|  |  | Gender attitudes | 1.7 |  |
|  |  | Power | 0.0 |  |
|  | None of the session topics disagreed with fathers' values or beliefs | Yes | 93.2 |  |
|  | Comfortability with mixed gender sessions | Very comfortable | 65.5 |  |
|  |  | Comfortable | 17.2 |  |
|  |  | Neutral | 13.8 |  |
|  |  | Uncomfortable | 1.7 |  |
|  |  | Very uncomfortable | 1.7 |  |
|  | Comfortability learning from a female delivery agent | Very comfortable | 29.3 |  |
|  |  | Comfortable | 53.4 |  |
|  |  | Neutral | 17.2 |  |
|  |  | Uncomfortable | 0.0 |  |
|  |  | Very uncomfortable | 0.0 |  |
|  | Comfortability learning from a male delivery agent | Very comfortable | 27.6 |  |
|  |  | Comfortable | 56.9 |  |
|  |  | Neutral | 15.5 |  |
|  |  | Uncomfortable | 0.0 |  |
|  |  | Very uncomfortable | 0.0 |  |

> *“Another thing I learned that I really liked was budgeting. Budgeting is something I really enjoyed, because before, I used to spend money in certain ways, but when they taught us how to budget, it really helped me a little Another thing that could bring benefits if we continue is budgeting, because as we know, many fathers can end up with money, but when they get home, they may find themselves spending it unwisely. However, after being taught budgeting, many of us have opened up and now understand how to manage our money better. It would have been better if this continued.”* (Father #7 at endline, Village #6)

Although we did not have quantitative data from mothers to directly compare, through the midline and endline qualitative interviews, most mothers commonly reported enjoying topics on nutrition, gender roles and attitudes, couples’ relationships, ECD, responsive caregiving, and playing with children.

> *“The topic that impressed me is listening parenting, because the baby has the right to be listened, when he needs something, you need to take care, you are not supposed to leave him alone. The other thing is child nutrition. I was not aware of how to prepare food for the baby, sometimes we were over cooking vegetables. Vegetables are warmed for five minutes, but you may find someone is over cooking, boiling, throw the eater and re boil then they eat, that is not supposed to be like that.”* (Mother #7 at endline, Village #6) *“For instance, the session on the relationship between husband and wife. It builds the family more than even childcare. A good relationship between husband and wife helps create a better family.”* (Mother #7 at midline, Village #5)

The session content was well received by caregivers with 71.0% of fathers and 77.8% of mothers reporting in the exit interviews that they shared information from the program with others in their communities. Additionally, in the endline quantitative survey, 96.5% of fathers reported sharing session content, and most often with friends (71%), neighbors (62%), and their partners (27%) to encourage positive behavior change within their households and communities (**Table 5**).

Nearly all fathers (93.2%) reported that none of the topics conflicted with their values or beliefs. However, among the small minority of fathers who expressed some reservations, responsive caregiving and positive discipline were the particular topics that were mentioned as raising some concerns. Qualitatively findings further revealed that positive discipline and shared household responsibilities were the relatively least acceptable topics, largely due to existing social and gender norms that traditionally defined parenting and domestic chores as a mother’s role and condone harsh punishment (i.e., hitting) as an acceptable form of discipline.

Nevertheless, some caregivers shared that with additional training, both for themselves and other members of their community, they would be more open to embracing these concepts.

> *“Ah, the topic that fathers should continue learning about, and that would be very helpful, is the one we just discussed about not hitting a child. Some of us were present, but we did not fully accept it. Some fathers, when they hear about it, might say, Oh, I cannot teach my child without using a stick. But if the teacher could continue developing that lesson, we would have learned more about how to calmly correct a child without always resorting to corporal punishment. They said, for me, my child would not understand without a little bit of discipline with a stick. I do not think they will understand if I just speak gently to them. But if the teacher had continued, we would have understood better how to handle such situations.”* (Father #7 at endline, Village #6) *“For example, men doing domestic chores, the things women are doing, they cannot do it… For example, if you ask a man to wash dishes, he cannot, they will say that you are stepping on his head It is very difficult in our community to change people, maybe for those who are trained.”* (Mother #7 at endline, Village #1)

#### Program materials

Fathers and mothers commonly described the full range of materials used in the program as helpful because they reflected their experiences, reinforced content, and offered clear, practical examples. Parents emphasized that the materials not only enhanced their engagement but also played a key role in fostering positive behavior change.

> *“It is good and practical. It doesn’t end there; there is a high possibility that they will continue playing at home. When we do it [playing with toys] during sessions it helps fathers who couldn’t play with their children learn from other fathers.”* (Father #1 at midline, Village #5)
>
> *“The pictures also are very helpful, they teach us how to live well in our families, they also teach about issues of violence in families and also knowing about stress management… They [the images] are educative and their messages really reflect the reality of what is happening in our communities.”* (Mother #14 at midline, Village #4)

Many fathers and mothers particularly valued the handouts because they served as a useful reference for key information discussed during the session. They reported storing the handouts distributed after each session, with a few specifically noting that they chose to hang them on a wall at home for easy access whenever they needed a reminder.

> *"Yes, they are important, they remind us of what we have been taught during the program… After two days, I look at them to remind myself of what I am supposed to do, like in dealing with stress, what am I supposed to do when dealing with stress? I read them and then select which one among them is best for me.”* (Father #7 at midline, Village #6)

Additionally, some fathers and mothers noted that the handouts facilitated knowledge sharing between partners, particularly when one had missed the previous session or when encouraging the other to participate. They reported learning about the session content by referring to the handouts brought home by their partners.

> *“Maybe the good thing is that even if you missed a session, there were handouts explaining the topics discussed. So, you could read and understand the basics. So, even if you were absent, it was easy to understand what was taught, although not as much as those who attended the sessions.”* (Father #7 at endline, Village #3)
>
> *“His work is limited, even if you say that you can change the time, still it will not work for him, though when I come I make sure I explain to him, and he agrees, that is why I was acting like his second teacher, when he sees the paper he reads it, he asks me questions and I explain to him.”* (Mother #7 at endline, Village #6)

Some caregivers also noted that the visual format of handouts and flipcharts made them accessible to individuals with low literacy or a hearing impairment. *“The use of pictures is good, like in our group there is one who has hearing impediment, she doesn’t hear clearly so the use of pictures is better than videos.”* (Father #7 at midline, Village #6) Similarly, many caregivers valued the videos, nothing that they clearly conveyed the key messages from sessions, accurately depicted their lived experiences, and helped reinforce the session content.

> *“The videos have been made with quality. When you learn and see the action then you have learnt twice. They should continue to create picture [the videos] because they add to our understanding. We are role playing so we can understand deeply. They are using big smartphones [to show the videos].”* (Father #11 at midline, Village #3)

#### Program model

The group sessions were conducted on a weekly basis, with each session lasting for about 2 hours. In the endline post-intervention survey 98% and 91% of fathers at reported that they liked this session format (*Table 5*). Similarly, the majority of both fathers and mothers interviewed at endline qualitatively expressed that they preferred meeting weekly. They stated that this session frequency allowed them to attend to other responsibilities while still giving them adequate time to apply and practice what they had learned between sessions.

The majority of caregivers preferred mixed-gender sessions over father-only or mother-only group sessions. Among exit interview respondents, 92% of mothers and 90% of fathers favored mixed-gender sessions. Additionally, in the endline quantitative survey with fathers, 83% reported being comfortable attending mixed-gender sessions, while 14% were neutral (**Table 5**).

Qualitative data at endline reinforced these findings, with most fathers and mothers expressing a clear preference for mixed-gender group sessions. They explained that attending together ensured both caregivers received the same information, which they believed would better encourage family-wide behavior change, and that mixed-gender sessions fostered more open and honest dialogue between partners.

> *“The lessons taught were for both men and women. If it’s violence, we are all affected, if it’s staying close with the family, we’re all in. We may have just a few that don’t concern both. They should be mixed because that’s how you build on a family. You cannot build a family when you are separated.”* (Mother #7 at endline, Village #5)
>
> *I felt good because we were all being taught together. Initially, when only we (the men) attended, we were the only ones learning. Our wives did not understand what was being taught. But later, we had very good sessions after being mixed with the women. It brought up discussions and questions during the sessions, which created a good learning environment.”* (Father #3 at endline, Village #3)

However, a few fathers and mothers noted that mixed-gender sessions with couples could sometimes make one parent less comfortable sharing openly in front of their partner. For example, one father preferred a blended approach, like how we had piloted, with mostly joint sessions supplemented by occasional father- or mother-only sessions.

> *“Yes, I suggest that some topics should be for fathers, for example, this week fathers come, and next week mothers come. To create a good image because there might be things I cannot talk about in front of my wife, or I could talk about them, but I don’t do so because my wife might oppose me in front of others. So, I believe that participating together is good, but also having sessions specifically for fathers only is important. That is, separate topics should be assigned that will address fathers, and certain topics should address mothers, but most of the topics should be for everyone together.”* (Father #7 at endline, Village #3)

#### Gender of facilitator

In addition to the sessions being primarily mixed gender, another gender-related aspect of the program was that the sessions were delivered by either a female or male facilitator. In the endline quantitative survey, none of the fathers reported feeling uncomfortable learning from a female or a male facilitator. The majority of fathers expressed feeling comfortable with both, with 83% feeling comfortable with the female facilitator and 85% with the male facilitator (**Table 5**), indicating no clear preference. This finding was further supported by qualitative data at endline, with both mothers and fathers stating that they were highly satisfied with both facilitators and irrespective of their gender.

## DISCUSSION

This pilot study provides promising preliminary evidence that *Familia Bora* is a feasible, acceptable, and appropriate parenting intervention for both mothers and fathers in a relatively low-income peri-urban setting in Mwanza, Tanzania. Participants expressed high levels of satisfaction across multiple domains of the program, including content, session activities, structure, materials, delivery modality, and facilitator approach. These findings were reinforced by diverse sources and methodologies, including in-depth interviews with mothers, fathers, group leaders, session observations, exit interviews, and facilitator feedback continuously over the course of program implementation. Our participatory, iterative, and evidence-informed co-design process that equally centered the experiences and perspectives of mothers and fathers, collaboration with a trusted local partner, and culturally relevant adaptations likely contributed to the program’s positive overall reception and engagement.

Program retention was encouraging, with low dropout and relatively high attendance rates. Overall, attendance rates averaged around 60% among men and 65% among women. The attendance rate for female caregivers in our pilot is comparable to that observed in other group-based parenting programs that have demonstrated meaningful impact [22, 23]. Notably, and unlike many prior interventions where fathers’ participation lagged far behind that of mothers [7, 15], we found near-equal attendance between men and women. This suggests that our intervention design effectively addressed barriers faced by not only women, but also men to foster conditions for more gender equitable participation in our program.

In addition to our co-design process grounded in formative research to ensure the content and delivery resonated with both mothers and fathers, we also tested a targeted implementation strategy where we provided travel allowance to encourage engagement. Our findings indicate that the travel reimbursements were well-received and played a key role in boosting attendance among both men and women. In a separate paper, we will explore this in greater detail, including a gender analysis comparing how financial incentives similarly or differently influenced mothers versus fathers. In the high poverty context of the study setting, the transport support was critical, echoing prior research that financial incentives can improve participation [24, 25], particularly by offsetting opportunity costs not only for men given prevailing gender norms, but also if not more for women who face considerable time and caregiving burdens as well. At the same time, it is important to carefully balance the use of transport allowance or financial incentives to optimize both engagement and long-term sustainability.

In addition to facilitators who led program delivery, we engaged community-based group leaders to support implementation by contacting participants, sending session reminders, and acting as liaisons between parents and the program. This strategy appeared effective in enhancing program feasibility by fostering participants’ sense of ownership and trust, while also assisting facilitators in managing logistical challenges. Evidence from peer-led and community-embedded models suggests that such roles can promote program uptake and responsiveness to community needs [26, 27]. However, we also discovered important challenges related to gender equity in this model. Initially, male group leaders were mostly focused on engaging other men, and women did not always receive direct reminders from the group leader. We identified this issue through midline data and took steps to course-correct by more actively and intentionally ensuring that women were also equally reached. Moving forward, we intend to have both male and female group leaders to ensure equitable outreach and support across genders.

In contrast to findings from other studies suggesting that men prefer sex-segregated sessions [28, 29], both fathers and mothers in *Familia Bora* overwhelmingly preferred mixed-gender groups. Even the few sessions initially designed to be for fathers only were met with requests from fathers to also include women so everyone could learn the same information and from each other. These findings suggest that preferences for gender-mixed or gender-separated sessions should be assessed within local contexts, as norms and desires may differ even within settings with restrictive gender expectations. Our results support prior studies from LMICs suggesting that couples parenting group sessions that incorporate opportunities for gender-specific activities or gender-separate discussions with also mixed-gender discussions can be both acceptable and effective [30].

Among all the topics covered, the sessions on couple relationships were the most enjoyed by men and women. These broadly included the sessions on conflict resolution, shared decision-making, partner support, and joint budgeting. Although lessons on positive couples’ relationship dynamics have been integrated in prior programs relating to family planning, violence prevention, and maternal health, they are less commonly emphasized in parenting programs focused on early child development. Our findings suggest that this content may not only improve family functioning but also serve as a compelling entry point for engaging both parents by focusing on their own relationship dynamics and needs, alongside promoting enhanced care for their child.

While most content was well-received, the one session topic that sparked some resistance was our content on discipline. Several participants expressed strong beliefs in the necessity of physical punishment, which clashed with the program’s emphasis on non-violent alternatives. While this tension is not entirely surprising considering sociocultural norms and high acceptability of corporal punishment in Tanzania and other similar settings [31-33], it underscores the need for program refinements to more effectively address these deeply rooted attitudes [34, 35]. In response, we are revising the content and activities to address these attitudes more directly by incorporating additional scenarios grounded in the local context, while demonstrating feasible, culturally appropriate alternatives to violence. This approach seeks to respect prevailing norms while providing a clear path toward positive behavior change, making the messaging more relatable, acceptable, and ultimately more effective in encouraging non-violent parenting practices.

Our use of multimedia tools – including flipchart images, videos, and the single-page handouts we distributed that included photos and the key message – proved to be a highly effective strategy for engaging parents and enhancing comprehension. Contextually tailored illustrations helped ground abstract concepts, and the visual materials ensured inclusive representation of both mothers and fathers. In particular, the videos helped parents better understand complex parenting behaviors, especially the importance of age-appropriate stimulation and responsive caregiving, which are topics that prior studies have identified as difficult to explain or demonstrate. Locally produced videos developed during the co-design phase effectively contextualized the content, featured both men and women, and were well-received by participants. While video-based approaches have been used in other global health domains, such as nutrition and health promotion [36, 37], they remain relatively underutilized in parenting interventions. Our findings, consistent with emerging evidence, suggest that incorporating multimedia may be especially effective in low-literacy settings, may offer additional pathways for scalable delivery, and warrants broader integration into parenting programs [38, 39].

A key strength of this study was its rigorous mixed-methods design, which enabled a comprehensive and nuanced understanding of program implementation and participant experience. We used both quantitative and qualitative methods and drew on multiple data sources from diverse perspectives. Our varied sampling strategy further strengthened the study as we used random sampling for baseline and endline qualitative interviews, purposive sampling at midline to include mothers and fathers with varying attendance levels, and random sampling for exit interviews to capture a broad range of experiences. This multi-method, multi-perspective approach enabled robust triangulation of findings, enhancing the validity of our findings and generating rich insights into the program’s acceptability, feasibility, and appropriateness.

Notwithstanding, the study has several limitations. The small sample size limits generalizability and limits the conclusions that can be drawn across other contexts and populations. The close involvement of M&E officers from the same implementing organization at every session may have introduced reporting or social desirability bias, potentially influencing participants’ responses. To mitigate this risk, we used multiple forms of data collection and included external research assistants to conduct midline and endline interviews, helping to reduce familiarity bias and encouraging more candid participant responses.

## CONCLUSIONS

This pilot study provides preliminary but compelling evidence that *Familia Bora* is a feasible, acceptable, and contextually appropriate intervention to support parenting in low-resource urban communities in Tanzania. By capturing multiple dimensions of participant experiences, program implementation, and contextual fit, the study offers valuable insights to guide further adjustments in program delivery and design and scale-up. We are currently making these adaptations and refinements through our research-practice partnership as we prepare for a cluster-randomized controlled trial. Future research directions to this work include the importance of assessing the program’s effects on paternal, maternal, and child outcomes, as well as its continued feasibility, fidelity, and acceptability, and overall implementation quality when integrated within community systems.

## Data Availability

All data produced in the present study are available upon reasonable request to the authors.

## ACKNOWLEDGEMENTS

We thank Amanda Dorsey for preparing an initial summary table of process indicators and Zane Maguet for assisting with qualitative analysis for part of the messages received section. We thank the research assistants who conducted the surveys and in-depth interviews and transcribed the qualitative data. Finally, we are deeply grateful to the mothers, fathers, and village leaders for their participation and support of our study. This study was funded by the Eunice Kennedy Shriver National Institute of Child Health and Human Development (R00HD105984).

